# Genetic patterns, sublineages and dynamics of tuberculosis transmission in Cuba, a low burden country

**DOI:** 10.1101/2024.03.04.24303662

**Authors:** Ileana Margarita Martinez-Rodriguez, Yoslany Mercedes Herrera-Avila, Roxana Goza-Valdes, Andrea Maria Rodriguez-Bertheau, Raul Diaz-Rodriguez

**Affiliations:** Medical Specialties Department. University of Medical Sciences-FAR, Havana, Cuba; Medical School. University of the Americas (UDLA). Quito, Ecuador; Microbiology Department. General Teaching Hospital ‘Enrique Cabrera’, Havana, Cuba; Medical Sciences School ‘General Calixto Garcia’. University of Medical Sciences-Havana, Havana, Cuba; National Reference and Research Laboratory of Tuberculosis, Leprosy and Mycobacteria. Institute of Tropical Medicine ‘Pedro Kouri’ (IPK), Havana, Cuba

**Keywords:** *Mycobacterium tuberculosis*, molecular typing, Cuba, tuberculosis, molecular biology, *Mycobacterium tuberculosis*/transmission, molecular epidemiology. molecular characterization

## Abstract

**Context:** In Cuba, a country with a low incidence of tuberculosis (TB), there is no information on the dynamics of transmission of the disease for two decades.

**Aims:** Determine the genetic patterns and sublineages of *Mycobacterium tuberculosis* complex (MTBC) isolates circulating in 2009 and their relationship with the epidemiological data of the patients.

**Settings and Design:** A retrospective descriptive study was carried out in Cuba with 178 MTBC isolates.

**Materials and methods:** Spoligotyping and mycobacterial interspersed repetitive-unit– variable-number tandem-repeat (MIRU-VNTR) typing with 24 loci were performed. For statistical analysis, non-parametric methods, analysis of variance and homogeneity test, were used.

**Results:** Spoligotyping produced 39 spoligotypes. The S, Beijing, LAM and Haarlem sublineages prevailed. The clustering rate was 75.84% and the Hunter-Gaston discriminatory index (HGDI): 0.8734. MIRU-VNTR typing with 24-loci defined 154 genetic patterns: six grouped 30 isolates and 148 showed unique patterns. The clustering rate was 14.60% and the HGDI: 0.9926. There was a predominance of sublineages by region: S, Beijing and LAM in the West, Center and East, respectively.

**Conclusions:** The dynamics of TB transmission in Cuba in 2009 is reported, inferring that this occurs in a limited manner and belonging to risk groups does not favor transmission. It serves to evaluate a megaproject of the Global Fund to reduce the transmission of TB in this country. The population genetic structure of MTBC resembles that of an Ibero-American country, with the exception of the high frequency of the Beijing and S sublineages.

## Introduction

Tuberculosis (TB) is an infectious disease considered a major public health problem.^[1]^ Molecular epidemiology, which emerged as a combination of molecular typing techniques for *Mycobacterium tuberculosis* complex (MTBC) and classical epidemiological approaches, has recently gained prominence as a resource for understanding crucial issues in the spread of TB.^[2]^

Adequate TB control requires knowledge of locally circulating isolates (or strains), the ability to distinguish between relapses and reinfections, identification of recent infections, associated risk factors, and the ability to track geographic distribution and clonal expansion of specific isolates or strains.^[2]^

The insidious forms of TB and the presence of undiscovered sources strongly support the use of typing methods in the effective control of epidemiological events, to identify points of infection in special situations or to search for the reasons for the spread of certain types of strains at the national. regional or global level. Interest in MTBC typing is further reinforced by the growing global threat posed by *M. tuberculosis* strains with multidrug resistance (MDR) and/or extensively drug resistance (XDR). ^[3]^

Cuba is one of the 15 countries with the lowest burden of the disease in the Americas region ^[4]^ and is working steadily to eliminate TB. In 2022, the WHO estimated a total incidence rate of 6.6 cases of TB per 100,000 inhabitants, with 83% of pulmonary TB being bacteriologically confirmed. ^[5]^ The rate in children/young people under 18 years of age was 1.0% and the estimated proportion of new cases with rifampin-resistant and multidrug-resistant TB (RR/MDR TB) was only 2.2%. ^[5]^ However, the incidence rate has remained around 6 per 100,000 inhabitants from 2004 to the present. ^[6]^

In Cuba, two population genetic studies were carried out in 1995 and 1998 using the restriction fragment length polymorphism analysis with the IS*6110* probe (IS*6110* RFLP), in which a clustering rate (reflecting recent transmission of the disease) of 48 and 45%, respectively, was found. ^[7, 8]^ In a molecular epidemiology study conducted by Gonzalez *et al*., using mycobacterial interspersed repetitive-unit–variable-number tandem-repeat (MIRU-VNTR) typing with 24 *loci*, on 61 MTBC isolates from 2009 in Havana, allowed corroborating that recent transmission was an important phenomenon in this place and that it was strongly associated to the permanence in closed institutions. It was also found that the conventional contact tracing in Havana fails to identify a relevant number of epidemiological links. ^[9]^ However, in Cuba, there is no information on the dynamics of TB transmission since the 1990s.

In 2009, a megaproject, financed by the Global Fund to fight aids, tuberculosis and malaria, began in Cuba (for five years) to strengthen the National TB Control Program (NTCP). It covered all components of the Program and included multiple sectors of Cuban society. However, the impact this had on reducing recent disease transmission and the variation of circulating genotypes is not known.

In the study presented here, we determined, using spoligotyping and 24-*locus* MIRU-VNTR typing, the genetic patterns and sublineages of MTBC isolates circulating in Cuba in 2009 and their relationship with the clinical-epidemiological data of the patients, and an estimate of the recent transmission of the disease, in order to establish a baseline.

## Materials and Methods

A retrospective analytical-descriptive study was carried out. The universe consisted of 666 TB cases diagnosed in 2009.

### Type of sampling and reasons for selection

Inclusion criteria: All Cuban MTBC isolates arriving at TB Lab in 2009 sample. Exclusion criteria: All repeated isolates, those that were contaminated or dried, those for which DNA was not obtained, or those for which molecular characterization was not completed were eliminated.

In the TB Lab, 383 MTBC isolates were received, obtained from clinical samples processed in the TB laboratories of the Provincial Centers of Hygiene, Epidemiology and Microbiology of the country. In total, 178 available DNAs (preserved at 4°C in sealed 1.5 mL plastic tubes) purified from the above MTBC isolates were used. This sample represented 26.72 % of the total number of patients reported and 46.47 % of the isolates received.

### Molecular typing

Spoligotyping was performed using the commercial system Kit IM9701 (Ocimum Biosolutions Ltd, Hyderabad, India). It was executed according to the methodology established by Kamerbeek *et al*. ^[10]^

MIRU-VNTR typing with 24 *loci* was performed according to the international protocol of Supply *et al*. ^[11]^

### Bioinformatic and statistical analysis

Genotypes obtained by 24-*locus* MIRU-VNTR typing (digital codes) and spoligotyping were analyzed with the online bioinformatics tools MIRU-VNTR*plus* (available at: https://www.miru-vntrplus.org/) ^[12]^ and SITVIT2 (available at: http://www.pasteur-guadeloupe.fr:8081/SITVIT2). ^[13]^

The definitions of clusters, unique or unclustered isolates and patients of a cluster proposed by Cave *et al*. (2005) were used. ^[14]^

The definition of clustering rate (or clustering percentage), as a minimum estimate of the proportion of TB cases related to recent transmission, as described by Small *et al*. ^[15]^ was used, according to the following formula: Number of patients in clusters-Number of clusters/ Total number of patients.

The following terms were also used in bioinformatics tools:

Allelic diversity: taking into account the values of genetic polymorphism of each *locus* or allelic diversity (h), highly (h> 0.6), moderately (0.3≤ h≤0.6) and poorly discriminatory (h< 0.3) *loci* were defined, according to Sola et al., 2003. ^[16]^

The discriminatory power of the typing techniques was determined by calculating the Hunter and Gaston Discriminatory Index (HGDI). ^[17]^

Sublineage: Group of isolates that share essential characteristics and descend from a common ancestor. ^[18]^

For the collection of clinical-epidemiological data on patients, the national TB database of the Ministry of Public Health (Minsap) for the aforementioned period and the database created in the TB Lab (and updated every year) with the results of the survey for the surveillance of resistance to antituberculosis drugs were reviewed.

In the statistical processing of the clinical-epidemiological data, the following non-parametric methods were used: analysis of variance and homogeneity test using the statistical package Statgraphics Plus version 2.1 (Statgraphics Technologies, Inc., The Plains, Virginia, USA). The results were expressed in the form of tables and figures.

### Ethical considerations

This study was approved by the IPK Ethics Committee (CEI-IPK-35-12), on April 20, 2012. It was conducted in accordance with the Declaration of Helsinki. All patients had a signed written consent previously approved by the ethics committee. They also discussed with a physician who clarified any doubts associated with the participation in the study. When patients were less than 18 years old an informed written and signed consent was obtained with the additional approval and sign of one of the parents. All sign consents were kept in physical files locked under the custody of principal investigators to maintain the anonymity of patients. Good laboratory practices were strictly followed, as well as all biosafety measures for the work and handling of microorganisms according to the risk levels established by the official list of biological agents that affect humans, animals and plants of resolution No 199/2020 of the Ministry of Science, Technology and Environment in force in Cuba.

## Results

Spoligotyping performed on 178 DNAs obtained from MTBC isolates from Cuba in 2009 yielded 39 different spoligotyping patterns. The calculated clustering rate was 75.84 % and the HGDI was 0.8734.

Four predominant sublineages of MTBC were identified, which included 84.26 % of the isolates studied. Individually, the S sublineage predominated (26.4 %), followed by the Beijing (23.5 %), LAM (19.66 %), and Haarlem (14.6 %) sublineages (Figure 1). MIRU-VNTR typing with 24 loci of the 178 MTBC isolates defined 154 different genetic patterns, of which six clustered 30 isolates (16.85 %) and 148 showed unique patterns, according to the online bioinformatics tool MIRU-VNTRplus (16). The clustering rate was 14.60 % and the HGDI was 0.9926.

**Figure 1.**
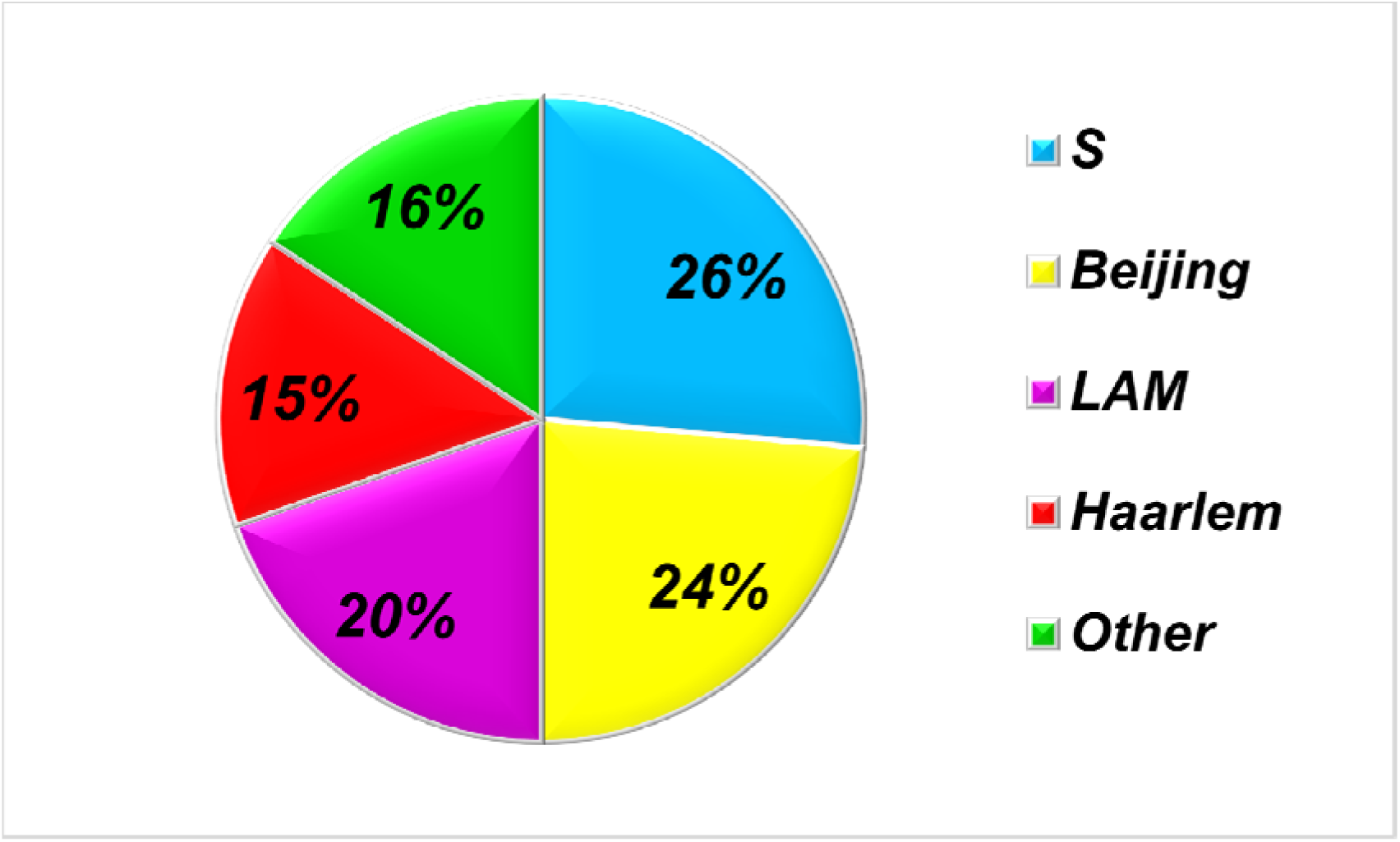
Main sublineages found in 178 isolates of the *Mycobacterium tuberculosis* complex obtained from TB patients in Cuba in 2009, according to the International Database SITVIT2 (Available in: http://www.pasteur-guadeloupe.fr:8081/SITVIT2).

Regarding allelic diversity (h), it was observed that of the 24 loci, nine were highly discriminative; ten moderately discriminative and finally there were 5 loci, poorly discriminatory (Figure 2).

**Figure 2.**
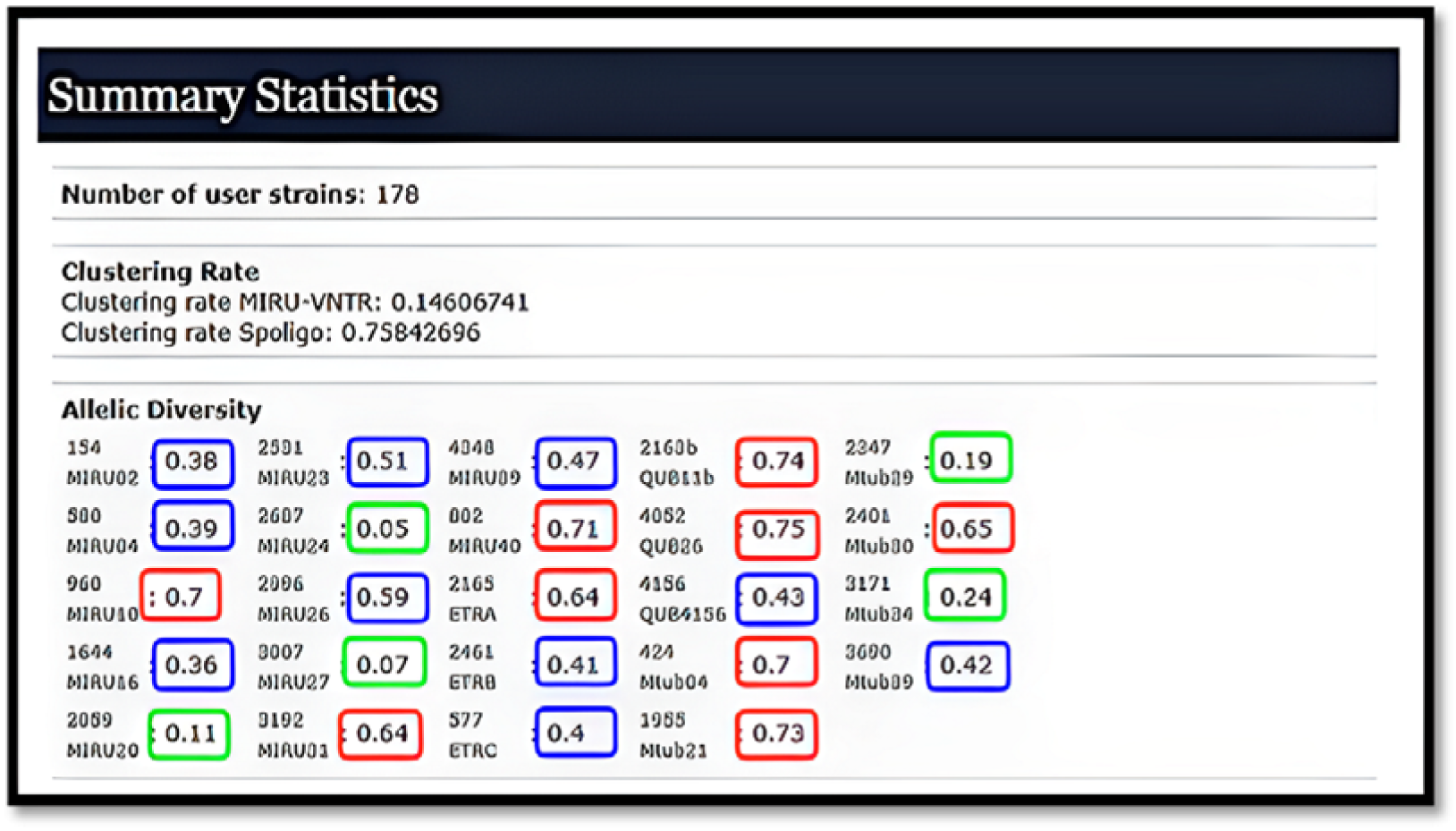
Result of the calculation of the allelic diversity of the 24-*locus* mycobacterial interspersed repetitive-unit–variable-number tandem-repeat (MIRU-VNTR) typing of 178 isolates of the *Mycobacterium tuberculosis* complex obtained in Cuba (2009), according to the online bioinformatic tool MIRU-VNTR *plus* (Available in: https://www.miru-vntrplus.org/).

Concerning the epidemiological information of the cases studied, it was observed that the mean age of the 178 patients was 43.19 ±15.18 years. The vast majority (91.57 %) of the cases were found in the age range of 15 to 64 years, mainly in the group of 30 to 44 years, and the remaining 8.42 % above 65 years. There was also a significant predominance of male cases (78.65 %). This proportion was maintained in both clustered and non-clustered patients (Table 1).

**Table 1.** Basic clinical-epidemiological characteristics of tuberculosis cases with *Mycobacterium tuberculosis* complex isolates obtained in Cuba in 2009, genotyped by 24-*locus* mycobacterial interspersed repetitive-unit–variable-number tandem-repeat (MIRU-VNTR) typing and Spoligotyping.

| Variable | Total | Non-clustered (148 cases) | Clustered (30 cases) |
| --- | --- | --- | --- |
| Sexo |  |  |  |
| Female | 38 | 31 | 7 |
| Male | 140 | 117 | 23 |
| Age Groups |  |  |  |
| 15 – 29 years | 41 | 33 | 8 |
| 30 – 44 years | 67 | 54 | 13 |
| 45 – 64 years | 55 | 47 | 8 |
| ≥65 years | 15 | 14 | 1 |
| Geographic origin |  |  |  |
| West | 89 | 66 | 23 |
| Center | 49 | 47 | 2 |
| East | 40 | 35 | 5 |
| Acid Fast Bacilli Smear |  |  |  |
| Positive | 154 | 128 | 26 |
| Negative | 24 | 20 | 4 |
| VIH Status |  |  |  |
| Positive | 25 | 18 | 7 |
| Negative | 153 | 130 | 23 |
| Resistance to antituberculous drugs |  |  |  |
| Resistant | 10 | 6 | 4 |
| Sensible | 168 | 142 | 26 |
| Permanence in closed institutions |  |  |  |
| Yes | 9 | 9 | 0 |
| No | 169 | 139 | 30 |
| Mortality |  |  |  |
| Yes | 14 | 10 | 4 |
| No | 164 | 138 | 26 |

When examined by region, it was observed that the West accumulated half of the cases (p=0.0465). When comparing the transmission dynamics by region it was distinguished that in the West there was a higher genetic diversity because a larger number of patterns were detected. Additionally, there was a higher recent transmission because there was a higher tendency to clustering (p=0.0034).

This difference in the behavior of the West is conditioned by the province of Havana City (named Havana province since 2011), which had the highest percentage of clustering (29.09%), while in the rest of the country, only 11.38% of the cases were found in clusters (Figure 3).

**Figure 3.**
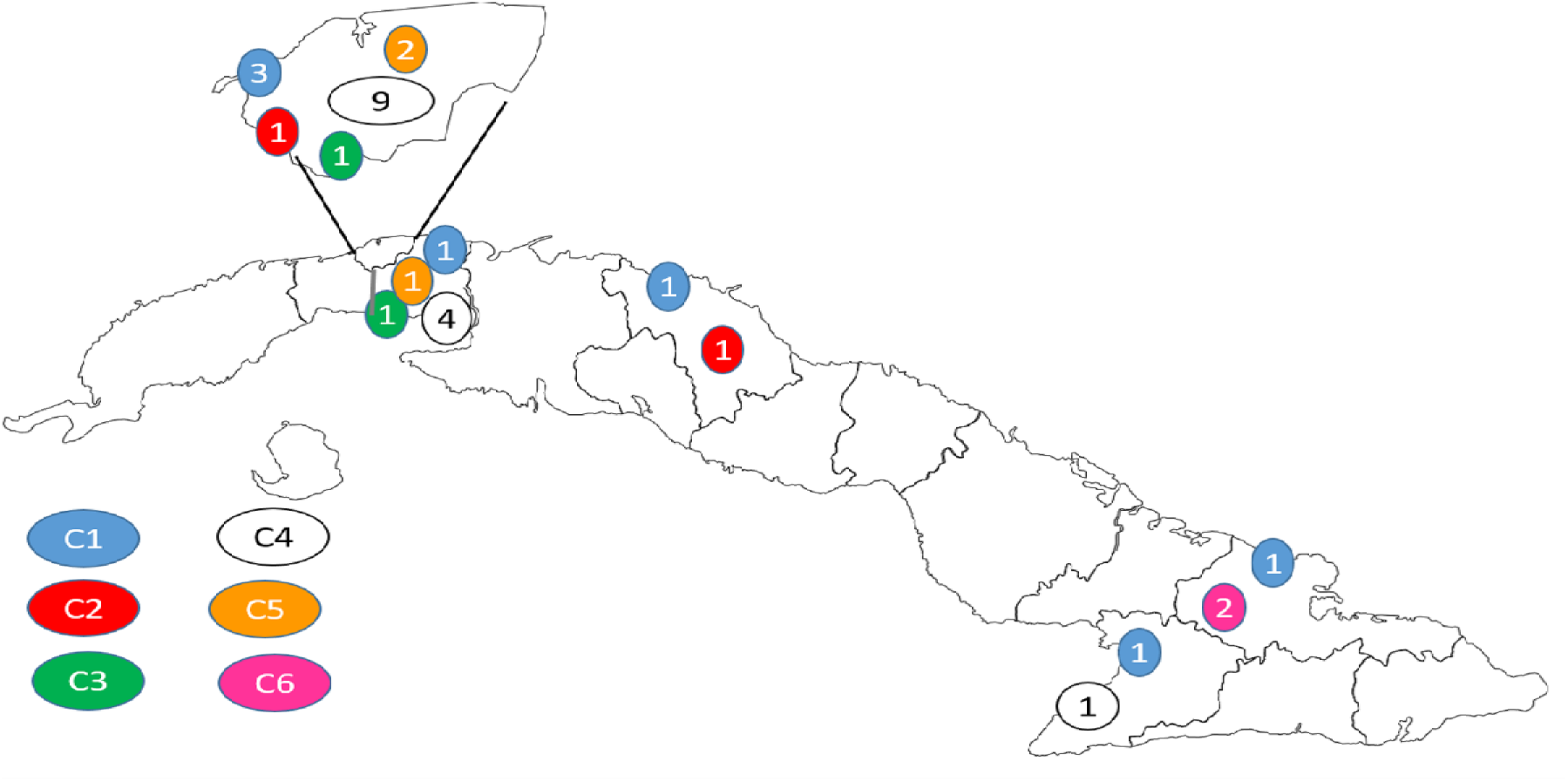
Geographical distribution of TB cases according to clusters found by 24-*locus* mycobacterial interspersed repetitive-unit–variable-number tandem-repeat (MIRU-VNTR) typing and provinces of Cuba.

When analyzing the distribution of sublineages by regions, a difference in distribution was observed (p=0.0087) with a different sublineage predominating in each one: the S sublineage in the West, Beijing in the Center of the country, and LAM in the Eastern region (Table 2). Regarding the distribution of the sublineages according to clusters, it was observed that 21.43 % of the isolates of the Beijing sublineage and 36.17 % of the isolates of S were found in clusters, in contrast to the other reported sublineages. However, there was no relationship between the Beijing sublineage and clusters (p =0.4496 with Yates correction) unlike S, where a strong association was observed between this sublineage and clusters (p =0.0001) (Table 3).

**Table 2.**
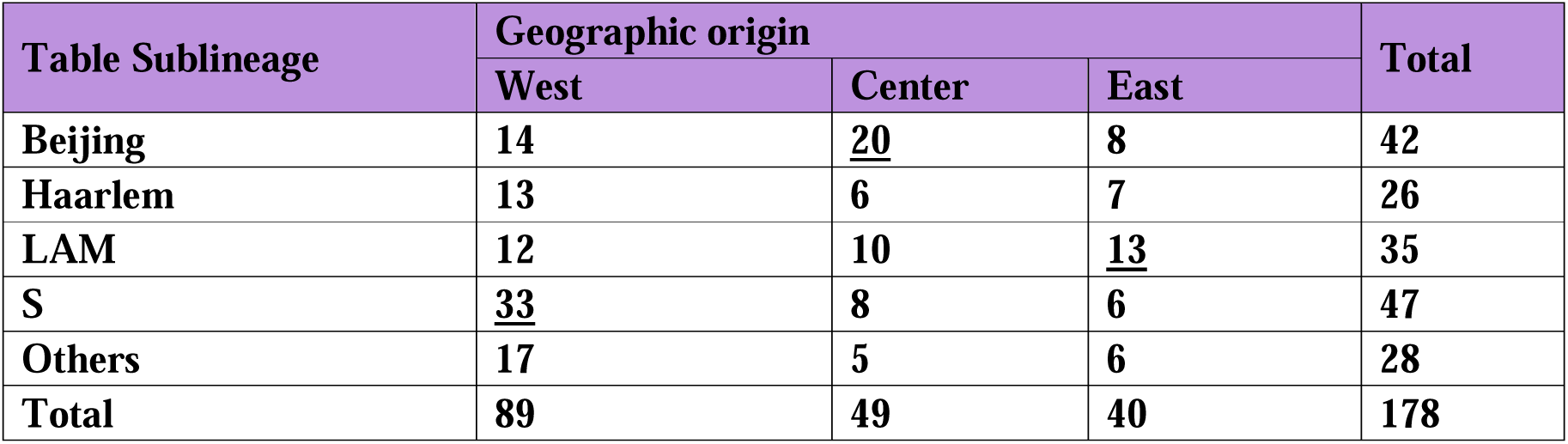
Geographical distribution of the sublineages of 178 *Mycobacterium tuberculosis* complex isolates obtained from TB patients of Cuba in 2009, according to the International Database SITVIT2 (Avaliable online: http://www.pasteur-guadeloupe.fr:8081/SITVIT2).

| Table Sublineage | Geographic origin |  |  | Total |
| --- | --- | --- | --- | --- |
|  | West | Center | East |  |
| Beijing | 14 | <u>20</u> | 8 | 42 |
| Haarlem | 13 | 6 | 7 | 26 |
| LAM | 12 | 10 | <u>13</u> | 35 |
| S | <u>33</u> | 8 | 6 | 47 |
| Others | 17 | 5 | 6 | 28 |
| Total | 89 | 49 | 40 | 178 |

**Table 3.** Distribution of the sublineages of 178 *Mycobacterium tuberculosis* complex isolates obtained from TB patients of Cuba in 2009, according to the International Database SITVIT2 (Available online: http://www.pasteur-guadeloupe.fr:8081/SITVIT2) and the clustering.

| Sublineage | Clustered | Non-clustered | Total |
| --- | --- | --- | --- |
| Beijing | 9 | 33 | 42 |
| Haarlem | 2 | 24 | 26 |
| LAM | 2 | 33 | 35 |
| S | 17 | 30 | 47 |
| Others | 0 | 28 | 28 |
| Total | 30 | 148 | 178 |

Only ten isolates resistant to at least one anti-tuberculosis drug were found, of which 40% were in clusters. The association between antituberculosis drug resistance and clustering was not well evidenced. Of these isolates, three were included in the first cluster by 24-locus MIRU-VNTR typing. This is the only one with a significant difference with respect to the rest of the studied isolates (p=0.0001). When analyzing the relationship between treatment resistance and sublineages, it was observed that, of the 10 cases, seven had MTBC isolates belonging to the Beijing sublineage, which evidenced a relationship between this sublineage and treatment resistance (p=0.0103) (Table 4).

**Table 4.** Relationship between the results of drug sensitivity tests on 178 *Mycobacterium tuberculosis* complex isolates obtained in Cuba in 2009 and sublineages, according to the International Database SITVIT2. (Available online: http://www.pasteur-guadeloupe.fr:8081/SITVIT2.

| Sublineage | Resistant | Sensible | Total |
| --- | --- | --- | --- |
| Beijing | 7 | 35 | 42 |
| Haarlem | 0 | 26 | 26 |
| LAM | 2 | 33 | 35 |
| S | 0 | 47 | 47 |
| Others | 1 | 27 | 28 |
| Total | 10 | 168 | 178 |

When studying some recognized risk factors, such as HIV infection or permanence in closed institutions, it was observed that of the 25 people living with HIV, only seven had isolates in the clusters and four of these were found in cluster C1. Although in general, there was no difference between clustered and non-clustered isolates (p=0.1876).

When associating the mortality of TB cases with the clusters determined with 24-locus MIRU-VNTR typing, it was observed that there were no differences between cases with MTBC isolates included in the clusters and those not included (p=0.3963). In relation to prolonged stay in closed institutions, it could be seen that all patterns were unique (Table 5).

**Table 5.** Distribution of 178 *Mycobacterium tuberculosis* complex isolates obtained in Cuba (2009) from patients included in clusters according to HIV status, permanence in closed settings, deceased.

| Variable | Percentage de presentation | Percentage of clustering | p Value |
| --- | --- | --- | --- |
| Deceased | 7,86 | 28,57 | p=0,3963 |
| permanence in closed settings | 5,05 | 0 | p=0,3528 |
| VIH Status | 14,04 | 28 | p=0,1876 |

## Discussion

In Cuba, since the 1990s, genotyping techniques (IS*6110* RFLP and spoligotyping) have been implemented for the molecular characterization of MTBC isolates obtained in this country and to carry out population-genetic studies. ^[7,8]^ With these studies, a clustering rate (reflecting recent transmission) of 48 and 45% was found in Cuba (1995) and Havana (1998), respectively.

In 2008, 15-*locus* MIRU-VNTR typing was introduced and later (2009) a molecular characterization study was carried out (with this technique) in 80 MTBC isolates obtained from patient samples from health units of Havana. A high clustering rate (53%) was found. ^[19]^

A more complete molecular epidemiological study was then carried out in this city (with MTBC isolates and TB patients from 2009) and it was corroborated that recent transmission was an important phenomenon in this place and that it was strongly associated to the permanence in closed institutions. It also shows that the conventional study of contacts in Havana fails to identify a relevant number of epidemiological links. ^[9]^ However, for all of Cuba, there is no information on the dynamics of TB transmission since the 1990s.

This work focuses on the genotypic study of MTBC isolates obtained in Cuba in 2009 and their interaction with related patients, the year in which a large five-year project began with support from the Global Fund to fight aids, tuberculosis and malaria to strengthen the PNCT. This has been the largest (US$7.9 million) and most comprehensive project (integrating all components of the Program and multiple sectors of Cuban society, including civil society) in the history of the PNCT in 60 years. Although a decade has passed since its completion, it is not known what impact it had on the reduction of recent transmission of the disease and on the variation of the circulating genotypes and sublineages, especially of some of them internationally notorious for their relationship with virulence, transmissibility and drug resistance, such as the Beijing sublineage. Therefore, the results of this work could serve as a baseline for understanding these aspects.

When analyzing the clustering rate of this work, it is similar to the research conducted by Pedersen *et al*. in the Nordic countries (Denmark, Sweden and Finland) in the periods 2012-2013 and 2014-2015. ^[20]^ It is lower than that found in Germany, Finland and the Netherlands, all countries with low TB incidence and effective control programs. ^[21–23]^

Rapid identification of the most discriminative *loci* saves time and resources during monitoring of isolates with particular characteristics, in field investigation of past or potential outbreaks, and tracking imported isolates. In comparison with a review and analysis study of 56 articles, published between 2002 and 2019, that evaluated the discriminatory power of each *locus*, it is observed that five of the six highly discriminative *loci* found in these studies correspond to those of this research. ^[24]^ A high concordance is also seen with the study conducted in Iran where six of the highly discriminative *loci* match. ^[25]^

The identification of the main lineages and sublineages and their geographical distribution allow a better understanding of the transmission dynamics of the disease. ^[13]^ The high frequency of occurrence of sublineage S is striking. This sublineage is also reported in France, Canada and South Africa, countries with which Cuba has increased trade relations in recent years. ^[13, 26]^ In Italy, it is considered an autochthonous family, as evidenced in a study conducted by Garzelli *et al*. in Tuscany where 92.5% of cases in native Italians belong to this sublineage. ^[27]^

The high proportion of isolates with LAM and Haarlem sublineages constitutes a reflection of the demographic origin of the Cuban population and the historical and persistent links with Latin America and Mediterranean Europe, regions in which these sublineages largely predominate. ^[13, 28]^

Although Cuba has one of the lowest TB rates in Latin America, it has a high prevalence of the Beijing sublineage, ^[7, 8]^ compared to other countries in the region, except Peru, Colombia and Ecuador. ^[29, 30]^ It is important to highlight that the presence of this sublineage in Cuba has increased from 11.3% to 25.6%, between 1995 and 2010, without knowing the possible causes. ^[7, 28]^

When analyzing the population included in this study, it is observed that the mean age of the population coincides with that found in the state of Veracruz, Mexico, and in the state of Romaira, Brazil. ^[31, 32]^ It differs from that reported in the foreign population in Finland where the mean age is 28 years and in Tokyo, Japan, where an age of less than 40 years is reported. ^[22, 33]^ It is evident that the mean age of the patients is lower as the incidence of the disease increases.

In this research, it was found that Havana City was the province with the most clusters, in addition to having the two largest groups in the study. This finding is similar to Izumi *et al*., who found that being a resident in cities was associated with clusters. ^[33]^

There is no association between HIV and clustering, which is due to the fact that HIV status favors the development of the disease, but not its transmission. This explains the similarity with the study conducted in 505 TB patients in Cape Town, South Africa. ^[34]^ Although there were differences in the incidence of TB, there were no differences between clustered and single cases.

TB mortality is more related to strain virulence and immunological characteristics of the patient than to recent transmission. The absence of association between clustering and mortality is similar to what was found in the study conducted in Malawi where there was also no association with mortality. ^[35]^ On the other hand, it differs from what was found in a study in Zimbabwe, where mortality was higher in clusters. ^[36]^

In this research, a very low percentage of resistance to antituberculosis treatment was observed, similar to the study conducted by Pedersen *et al*., in Northern European countries, ^[18]^ and contrarily to investigations in some Asian or African countries, such as India and Mali. ^[37, 38]^ This low incidence of drug resistance may be due to the adequate management of patient treatment and the low importation of resistant cases. The non-association between resistance and clusters may be due to the low occurrence of isolates resistant to antituberculosis drugs. In conclusion, it can be said that this research contributes to the knowledge of the recent transmission dynamics of TB in Cuba in 2009. The size of the clusters and the high number of unique patterns allows us to infer that TB in this country occurs limited form and belonging to risk groups does not favor the transmission of the disease.

Additionally, it serves as a benchmark to help assess the positive impact that the financing of the Global Fund megaproject may have had on reducing the spread of MTBC isolates in the community and decreasing TB transmission in this country in subsequent years.

The genetic population structure of MTBC found in Cuba resembles that of an Ibero-American country, with the exception of the high frequency of appearance of the Beijing and S sublineages. ^[29, 30]^

In order to update knowledge on the recent transmission of the disease in Cuba, a molecular typing study with MTBC isolates obtained in 2017-2019, prior to the negative influence of the COVID-19 epidemic in the country, is underway.

### Limitation of study

This study has some limitations. i) Not all *M. tuberculosis* isolates cultured during the period were analysed, as some isolates (or DNAs) were lost (or without fully molecular results) during storage, transportation, DNA extraction, spoligotyping or 24-*loci*-MIRU-VNTR typing. i) the time period was the only one year, missing clustered isolates from linked patients in recent transmission events out of this short period.

## Data Availability

All data produced in the present study are available upon reasonable request to the authors

## Acknowledgements

The authors would like to thank:

– All the members of the Cuban National Network of TB Laboratories for their contribution in processing clinical samples and sending MTBC isolates, essential for DNA extraction.
– The staff of the National Reference Laboratory of TB of the IPK for the excellent technical assistance and the facilities guaranteed for the successful execution of this work.
– Dr. Howard Takiff, Dr. Maria Victoria Mendez, Arnout Mulder and Dr. Jessica de Beer for facilitating training in 24-*locus* MIRU-VNTR typing.
– To Dr. Annelis Bunschoten and Dr. Sofia Samper for helping in the training in Spoligotyping.

The authors carried out this research with the support of the following international projects:

– ’Development of immunodiagnosis methods, resistance detection, molecular characterization and implementation of epidemiological surveillance systems for tuberculosis control’ Supported by Misión Ciencia. Cuba-Venezuela Cooperation. (2008-2012).
– ’Strengthening the Tuberculosis Program in the Republic of Cuba’. Funding: Global Fund to Fight AIDS, Tuberculosis and Malaria (2009-2013).
– ’Strengthening research capacities, knowledge and actions to accelerate progress towards the elimination of tuberculosis in Cuba’ (FA4). Financial supported by Directorate-General for International Cooperation and Development (DGD), Belgium (2017-2021).

## Author Contributions

IMM-R and RD-R planned and made concepts, design, definition of intellectual content. IM M-R, YMH-A, RGV and RD-R executed the investigation. IMM-R and AMR-B performed curation and validation of data. IMM-R, AMR-B and RD-R wrote the first draft of the manuscript. All authors reviewed, editing and approved the final manuscript.

